# Synchronised inflations generate greater gravity dependent lung ventilation in neonates

**DOI:** 10.1101/2020.06.12.20129379

**Authors:** G Dowse, E Perkins, J Thomson, N Schinckel, PM Pereira-Fantini, DG Tingay

**Affiliations:** Neonatal Research, Murdoch Children’s Research Institute, Parkville, Australia; Department of Paediatrics, The University of Melbourne, Parkville, Australia; Department of Neonatology, The Royal Children’s Hospital, Parkville, Australia

**Keywords:** Synchronisation, Infant, Regional ventilation, mechanical ventilation

## Abstract

**Objective:** Synchronising positive pressure inflations with an infant’s own breathing is considered lung protective, but the ventilation patterns within the infant lung during synchronous and asynchronous inflations are unknown. The aim of this study was to describe the regional distribution patterns of tidal ventilation within the lung during mechanical ventilation that is synchronous or asynchronous with an infant’s own breathing effort.

**Methods:** Intubated infants receiving synchronised mechanical ventilation at The Royal Children’s Hospital NICU were studied. During four 10-minute periods of routine care, regional distribution of tidal volume (V_T_; Electrical Impedance Tomography), delivered pressure and airway flow (Florian Respiratory Monitor) were measured for every inflation. Post hoc, each inflation was then classified as synchronous or asynchronous from video data of the ventilator screen, and the distribution of absolute V_T_ and delivered ventilation characteristics determined.

**Results:** 2749 inflations (2462 synchronous) were analysed in 19 infants; mean (SD) age 28 (30) day, GA 35 (5) weeks. Synchronous inflations were associated with a shorter respiratory cycle (p=0.004) and more homogenous V_T_ (centre of ventilation) along the right (0%) to left (100%) lung plane; 45.3 (8.6)% vs 48.8 (9.4)% (uniform ventilation 46%). The gravity dependent centre of ventilation was a mean (95% CI) 2.1 (−0.5, 4.6)% more towards the dependent lung during synchronous inflations. Tidal ventilation relative to anatomical lung size was more homogenous during synchronised inflations in the dependent lung.

**Conclusion:** Synchronous mechanical ventilator lung inflations generate more gravity dependent lung ventilation and more uniform right to left ventilation than asynchronous inflations.

## INTRODUCTION

Mechanical ventilation is a common and life-saving therapy for many newborn infants needing intensive care. However, even brief periods of ventilation when applied inappropriately can injure the underdeveloped lung and may ultimately result in the development of chronic lung disease.[1-3] The initiation of lung injury is complex, often caused by opposing mechanisms such as atelectasis and volutrauma. As such there is increasing awareness of the need to ensure pressure and tidal volume (V_T_) are delivered evenly (homogenously) throughout the lung.[4-7] Despite this, reducing ventilation heterogeneity as a goal of lung-protective ventilation is rarely described in infants.[3,8,9]

Synchronising positive pressure inflations with the infant’s own inspiratory effort is advocated as a method of both improving patient comfort and lung protection, and hence is the most commonly used advanced ventilation mode in the Neonatal Intensive Care Unit (NICU).[10] Randomised controlled trials in preterm infants have shown that synchronisation improves oxygenation,[11] whilst reducing the delivered positive inflating pressure (PIP),[11-13] work of breathing[14] and the duration of mechanical ventilation.[11,15-17] Despite the short-term benefits of synchronised ventilation, a reduction in chronic lung disease has not been reported in preterm infants.[13]

Key to addressing the disparity between short and long-term outcomes is the development of an understanding of the impact of synchronous inflations on the V_T_ and flow patterns throughout the infant lung. Synchronisation promotes laminar gas flow within the lung, which may be associated with a more homogenous distribution of gas and therefore more protective than chaotic flow.[13] However, no studies to date have described the regional ventilation patterns during synchronous ventilation. Until recently this was due to an inability to measure regional flow and volume characteristics within the lung on a breath-to-breath bases. Neonatal specific electrical impedance tomography (EIT) systems now allow for the non-invasive, radiation-free measurement of regional changes in ventilation in the lungs of infants.[18] The hypothesis that synchronised inflations create a unique pattern of ventilation homogeneity can be tested for the first time.

This prospective, observational study aimed to describe the regional distribution of tidal ventilation within the lung during mechanical ventilation that is synchronous or asynchronous with an infant’s own inspiratory effort. Specific aims included defining the breath-to-breath characteristics of synchronous and asynchronous inflations within the right and left lung and between gravity dependent and non-dependent lung regions.

## METHODS

This study was performed in the NICU of The Royal Children’s Hospital (Melbourne, Australia), a quaternary referral unit managing both surgical and medical neonatal conditions, between March and September 2018. The study was approved by the institution’s Human Research Ethics Committee (HREC 36159A). Written, informed, parental consent was obtained for each infant.

Stable, intubated infants were eligible if they were less than 44 weeks corrected post-menstrual age at the time of study and receiving any mode of synchronised ventilation, with or without volume targeting (SLE5000, South Croydon, UK; a dedicated infant ventilator that synchronises inflations via a hot-wire anemometer at the airway opening). There were no restrictions to inclusion based on indication for ventilation, respiratory disease state or gestational age. Infants who were unstable during handling, had any condition that would alter the interpretability of EIT data (such as congenital diaphragmatic hernia), or if EIT belt placement interfered with medical care were not studied.

Infants were studied supine whilst settled. The regional distribution of V_T_ (V_TEIT_) was measured using the Pioneer EIT system with a non-adhesive 32 electrode neonatal EIT belt (SenTec AG, Landquart, Switzerland), as we have described previously.[19,20,21] Flow, pressure and V_T_ at the airway opening (V_Tao_) were recorded using the Florian Respiratory Function Monitor (Acutronic, Hirzel, Switzerland) integrated into the ventilator circuit during nursing care. A webcam was used to film the ventilator screen to allow later identification of inflations as synchronous or not. Four 10-minute continuous recordings were taken over approximately one hour at the ventilator parameters set by the clinical team. Florian and webcam data were measured simultaneously using LabChart (AD Instruments, Sydney, Australia) sampling at 200Hz and EIT data were imaged at 48 frames/sec using the manufacturer’s image acquisition software. Physiological data including peripheral capillary oxygen saturation (SpO_2_), heart rate, respiratory rate, blood pressure and end-tidal CO_2_ were manually recorded from the bedside Intellivue monitor MP70 (Philips Healthcare, Eindhoven, Netherlands) at one-minute intervals.

EIT images were reconstructed to create images of the distribution of air within the lung tissue and analysed using the manufacturer’s software package (ibeX, SenTec AG), in accordance with standardised guidelines.[22,23] From each 10-minute recording, one-minute of stable, artefact-free data were selected. EIT measured inflations were matched to Florian measured inflations, coded as synchronous or asynchronous and artefacts were excluded. A synchronous inflation was defined as a delivered PIP initiated by the infant’s inspiratory gas flow exceeding the set trigger sensitivity (the ventilator algorithm also code spontaneous effort with an orange colour change in the pressure and flow waves display).[12] An asynchronous inflation was defined as a delivered PIP without evidence of infant-generated inspiratory flow. Spontaneous inflations that occurred out of phase with the onset of PIP were excluded. In the case of modes using time and flow-cycled ventilation, only time-cycled inflations were included. These data could not be electronically outputted, so a webcam was used to film the ventilator screen, and each potential inflation for inclusion assessed by one researcher (GD). For each included inflation, the V_Tao_, PIP, positive end-expiratory pressure (PEEP), change in pressure (ΔP; PIP-PEEP), inspiratory time and respiratory cycle duration were calculated from the Florian data.

Functional EIT images of the distribution of V_TEIT_ for each coded inflation were created (Figure 1A) and data were extracted from the whole lung, right and left lung, gravity dependent (dorsal) and non-dependent (ventral) lung and the lung in three gravity dependent (ventral, central, dorsal) regions of interest (ROIs).[7,18,19,24,25] From these, standardised EIT measures to describe ventilation homogeneity were calculated: 1) the centre of ventilation (geometric mean of V_TEIT_ distribution along a single plane; Figure 1B) along the right-left (CoV_RL_) and 2) gravity non-dependent to dependent plane (CoV_VD_), and 3) the percentage of unventilated lung tissue.[18] To account for differences in the anatomical distribution of lung tissue within the chest, the V_TEIT_ homogeneity ratio (the ratio of the percentage of total lung V_TEIT_ delivered to a region to the percentage of total lung tissue contained within that region) was calculated in the three gravity-dependent ROIs (Figure 2A). A value of <1 suggests a relatively under-ventilated lung and >1 over-ventilated. Investigators were not blinded to the inflation type during analysis, but used in-built unmodifiable EIT algorithms in ibeX to calculate all ventilation variables.

**Figure 1.**
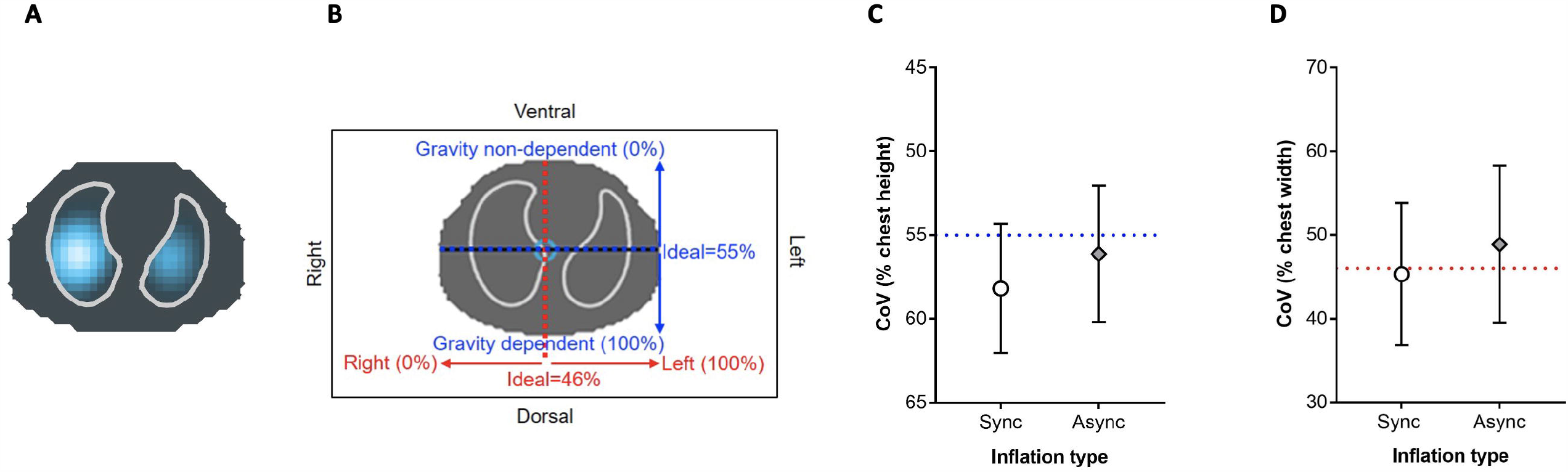
**A.** Representative functional EIT image of the distribution of V_TEIT_ during a synchronous inflation showing greater V_TEIT_ in the dependent lung than non-dependent, and a predominance overall of more V_TEIT_ in the central regions. Relative V_TEIT_ within the lung displayed using a colour scale with greatest V_T_ in white and least in dark blue. **B**. Schematic showing the ideal CoV when ventilation is homogenous. Along the right (0%) to left (100%) plane (red) ideal CoV is 46%.[18,22,23] Along the gravity non-dependent (0%) to gravity dependent (100%) plane (blue) ideal CoV is 55%.[18,22,23] **C**. Gravity dependent CoV for synchronous (Sync; open circles) and asynchronous (Async; grey diamonds) inflations compared to the uniform CoV of 55% (blue line). **D**. CoV along the right to left lung plane (uniform CoV 46%; red line) for pooled synchronous and asynchronous inflations using the same symbols as Panel C. Mean ± SD. Welch’s t-test with cluster analysis.

**Figure 2.**
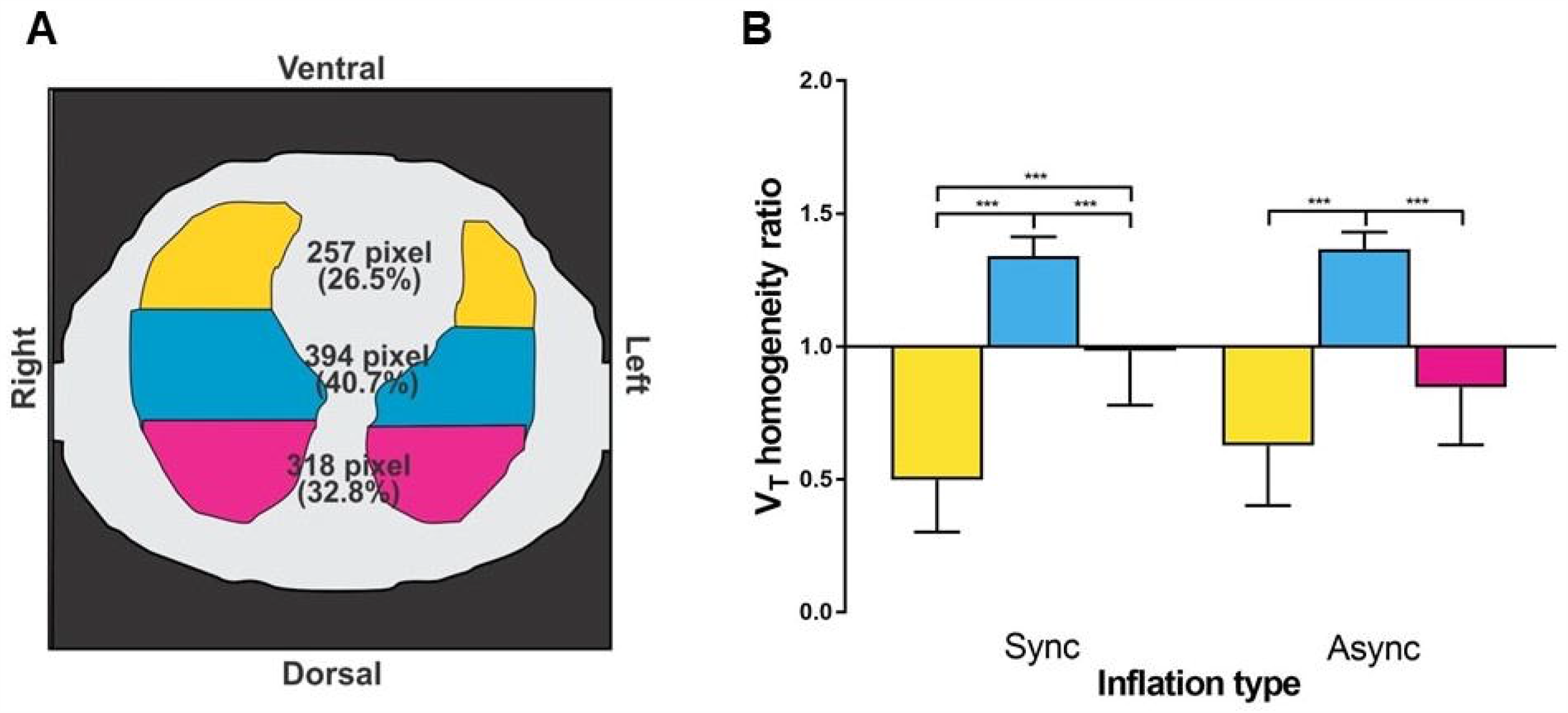
**A.** Schematic depicting the number of pixels, and anatomical percentage of total lung tissue, within the gravity non-dependent (yellow), central (blue) and gravity dependent (pink) regions of interest. **B**. Ratio of the measured percentage of total V_TEIT_ to the anatomical percentage of lung tissue within each third for the 2462 synchronous (Sync) and 287 asynchronous (Async) inflations using the same colours as Panel A. Mean ± standard deviation (SD). ***p<0.0001 (mixed effects with cluster analysis).

### Statistical analysis

A convenience sample size of 21 infants was chosen to generate 100-200 inflations/infant after outlier removal (ROUT method; Q=1%) for evaluation. Based on previous infant studies of EIT, a difference in CoV_VD_ of 2% would represent a significant change in V_T_ heterogeneity, and require 18 infants (assuming a 3% SD, power 80% and alpha error 0.05). Continuous outcomes between synchronous and asynchronous inflations were compared with a Welch’s t-test or mixed effects as appropriate, with robust standard error (cluster analysis) to account for multiple inflations from each infant. Linear regression was used to assess the relationship between important confounders that may have independently influenced regional ventilation characteristics irrespective of synchrony; gestational age (GA) at birth, age at the time of study and oxygenation deficit, although the study was not powered specifically for these associations. Statistical analysis was performed using Prism (v7.0, GraphPad Software Inc., California, USA) or STATA (v15.0, Statacorp, Texas, USA) and a p value <0.05 considered significant.

## RESULTS

Data from 19 of 21 infants were available for analysis (Supplementary Figure 1). Demographic characteristics are summarised in Table 1. Fourteen infants were receiving synchronised intermittent positive pressure ventilation at time of study, and five synchronised mandatory intermittent ventilation (one with additional pressure support). Sixteen infants were managed with volume targeted ventilation. Trigger sensitivity ranged between 0.2-0.6 L/min. A total of 2749 ventilator-delivered inflations were analysed; 2462 synchronous (90%) and 287 asynchronous (10%).

**Table 1.**
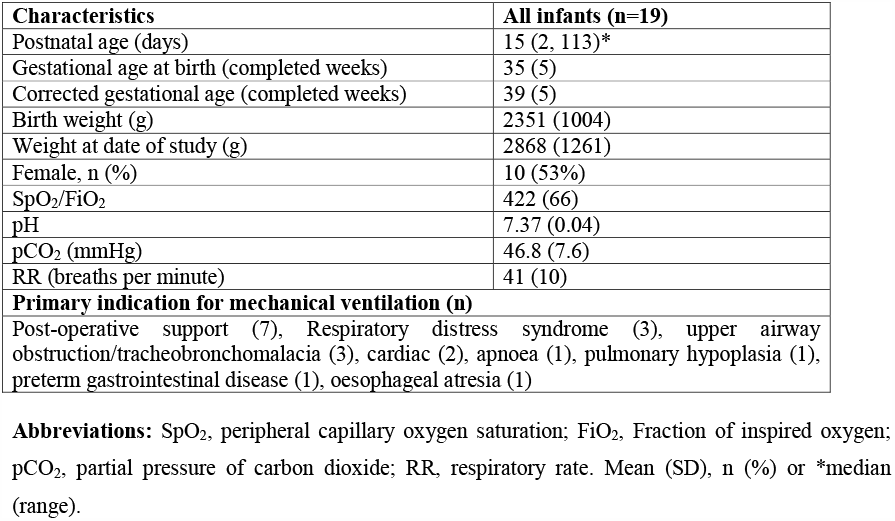
Infant characteristics.

| **Characteristics** | **All infants (n=19)** |
| --- | --- |
| Postnatal age (days) | 15 (2, 113)* |
| Gestational age at birth (completed weeks) | 35 (5) |
| Corrected gestational age (completed weeks) | 39 (5) |
| Birth weight (g) | 2351 (1004) |
| Weight at date of study (g) | 2868 (1261) |
| Female, n (%) | 10 (53%) |
| SpO<sub>2</sub>/FiO<sub>2</sub> | 422 (66) |
| pH | 7.37 (0.04) |
| pCO<sub>2</sub> (mmHg) | 46.8 (7.6) |
| RR (breaths per minute) | 41 (10) |
| **Primary indication for mechanical ventilation (n)** |  |
| Post-operative support (7), Respiratory distress syndrome (3), upper airway obstruction/tracheobronchomalacia (3), cardiac (2), apnoea (1), pulmonary hypoplasia (1), preterm gastrointestinal disease (1), oesophageal atresia (1) |  |

**Abbreviations:** SpO<sub>2</sub>, peripheral capillary oxygen saturation; FiO<sub>2</sub>, Fraction of inspired oxygen; pCO<sub>2</sub>, partial pressure of carbon dioxide; RR, respiratory rate. Mean (SD), n (%) or \*median (range).

### Delivered pressure and volume characteristics

Synchronous inflations were associated with a shorter respiratory cycle and inspiratory time than asynchronous inflations (Table 2). There was no difference in PEEP, V_Tao_ and ΔP between synchronous and asynchronous inflations, with ΔP being <6 cm H_2_O on average overall. Individual infant data for each outcome are available in Supplementary Figures 2-6.

**Table 2.**
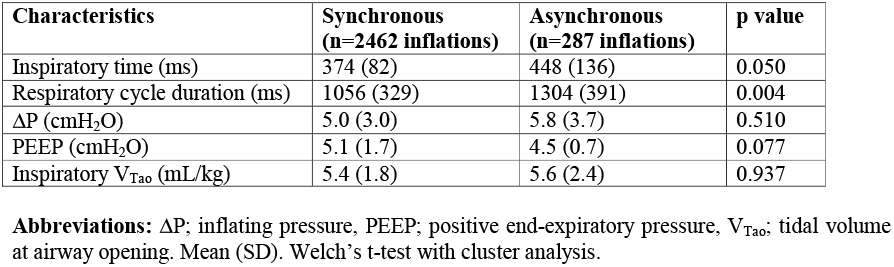
Pressure and volume characteristics during synchronous and asynchronous inflations.

### Regional distribution of tidal ventilation

The distribution of V_TEIT_ was highly variable within and between infants for both synchronous and asynchronous inflations (Supplementary Figures 7-10). Overall, ventilation favoured the gravity dependent hemithorax during both inflation types (Figure 1C). CoV_VD_ was a mean (95% CI) 2.1 (−0.5, 4.6)% towards the dependent lung during synchronous inflations than asynchronous (Welch’s t-test with cluster analysis). CoV_RL_ was more homogenous during synchronous than asynchronous inflations; mean (SD) 45.3 (8.6)% and 48.8 (9.4)% respectively, mean difference 3.6 (0.0, 7.2)% (Figure 1D).

During both inflation types, the central third was relatively over-ventilated compared to the gravity non-dependent and dependent regions (Figure 2B; p<0.0001; mixed effects models). The V_TEIT_ homogeneity ratio within the gravity non-dependent and dependent regions was not different between synchronous and asynchronous inflations (Figure 2B; both regions p=0.06). The non-dependent lung was under-ventilated during both inflation types. The dependent lung was also under-ventilated during asynchronous inflations, whilst synchronous inflations resulted in ideal ventilation.

### Percentage of unventilated lung tissue

There was no difference in the percentage of unventilated lung tissue during synchronous and asynchronous inflations, both approximating 8% of the lung regions (Supplementary Table 1). There was a mean (95% CI) 4.1 (2.6, 5.5)% more unventilated lung tissue within the non-dependent lung compared to dependent during synchronous inflations.

### The effect of infant characteristics on regional ventilation patterns

Increasing GA at birth and improving oxygenation (increased SpO_2_/FiO_2_ ratio) were associated with more homogenous gravity dependent ventilation, whilst increasing age at time of study resulted in greater gravity dependent lung ventilation (Figure 3). There was no relationship between CoV_RL_ and GA (p=0.58), age (p=0.83) or SpO_2_/FiO_2_ ratio (p=0.95, linear regression). These relationships were independent of synchronicity of ventilation.

**Figure 3.**
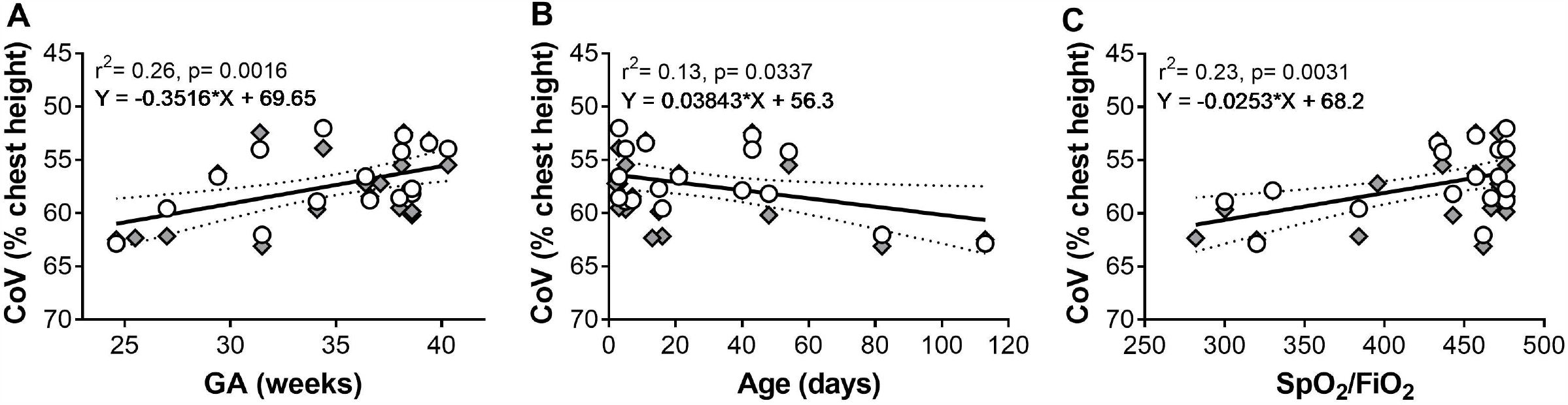
Relationship between gravity dependent CoV (%) and gestational age (GA) at birth (**A**), age at time of study (**B**) and SpO_2_/FiO_2_ ratio (**C**) for pooled synchronous (open circles) and asynchronous (grey diamonds) inflations. More mature GA at birth (p=0.0016, r^2^ 0.26), younger age at time of study (p=0.0337, r^2^ 0.13) and better oxygenation (p=0.0031 r^2^ 0.23) were associated with increasing non-dependent CoV (linear regression). Lines of best fit (95% CI dashed lines).

## DISCUSSION

The purpose of this observational study was to determine whether synchronisation improved homogeneity of ventilation throughout the lung; a proposed but not proven lung protective mechanism of synchronisation. We identified that ventilation homogeneity was highly variable during both synchronous and asynchronous inflations in stable, intubated infants, and overall ventilation was preferentially distributed towards the gravity dependent lung. This was more pronounced during synchronous inflations. Gravity dependent regional ventilation was independently influenced by birth GA, age and oxygenation. In addition, synchronous inflations generated a more homogenous right to left distribution of tidal ventilation.

In intubated infants with acute lung disease, tidal ventilation favours the non-dependent lung due to the effects of gravity on atelectasis.[24-27] Dependent regions are harder to inflate due to higher opening pressures imposed by less dependent lung tissue.[26,27] We did not find this pattern in our diverse population of stable infants, many without active lung disease. CoV_VD_ was similar to the preferential dependent lung ventilation that is reported in healthy, spontaneously breathing, term infants[28-31] and adults.[32] This pattern is attributed to the effect of gravity decreasing resting lung volume. The dependent lung therefore has greater capacity to expand to larger volumes during inspiration and does so with mechanical efficiency due to the role of negative pressure flow gradients and lack of disease.[28] The CoV_VD_ difference of 2.1% between synchronous and asynchronous inflations was not statistically significant due to the use of robust standard error analysis but is clinically relevant,[5] and likely reflects the greater contraction of the diaphragm in the dorsal chest during inspiration.[28,30] The posterior fibres of the diaphragm shorten the most, which increases dorsal lung volume.[28] The uniformity of tidal ventilation relative to anatomical size in the dorsal lung during synchronous inflations supports this. Further investigation using enhanced EIT measures which assess regional intra-tidal filling characteristics for homogeneity [33,34] may elucidate whether the distribution of ventilation to dependent regions during synchronous inflations is physiologically abnormal or beneficial.

During both synchronous and asynchronous inflations, central lung regions received the greatest V_TEIT_ relative to anatomical size, consistent with previous regional ventilation studies.[24,25,28] This could increase the risk of volutrauma; however, the central lung regions include the major airways and adjacent alveoli inflate more easily. EIT is unable to delineate between airways and alveoli. The finding of relative under-ventilation in both the gravity non-dependent and dependent ROIs during asynchronous inflations suggests greater potential for atelectasis in those lung regions. Although our study did not measure injury, the finding of regional differences in under and over-ventilated lung regions suggests that states of atelectasis and volutrauma can be created within the lung at the same time.[35-37] This complexity of injury potential has not been measured to date, and is not captured in current definitions of long-term lung injury outcomes.

An interesting finding was the high percentage and variability (0-20%) of unventilated non-dependent lung, especially during synchronous inflations. There are no previous reports of breath-to-breath variability of unventilated lung tissue. It was not surprising that unventilated lung tissue was greater in the distal lung. These regions are furthest from central airways, hardest to ventilate, and also at the boundaries of the image reconstruction models and may contain non-lung tissue.[18] The classical understanding of neonatal lung injury is that atelectasis is harmful,[2,3] but this concept is built on a general and persistent pattern of atelectasis within a lung region. Very little is understood about the role of variable atelectasis within alveoli, except that it can only be ameliorated with sufficient PEEP.[38] A more detailed examination of the unventilated lung patterns with new dynamic EIT measures of regional flow characteristics may identify new clinical approaches to measuring and managing atelectasis.

Right versus left lung ventilation homogeneity was more likely during synchronous inflations. Classical respiratory physiology dogma proposes the right lung of spontaneously breathing infants is easier to ventilate due to lower resistance in the shorter, narrower and more vertical right main bronchus.[29] As such, the left lung dominance during asynchronous inflations was unexpected. The shorter Ti during synchronous inflations is likely due to the faster gas flow rates from the combined contribution of the ventilator (positive pressure) and diaphragm (negative pressure), allowing targeted tidal volumes to be obtained quicker. Synchronisation is known to generate more laminar flow.[13] This is likely to distribute gas more uniformly than the more chaotic flow patterns during asynchronous inflations, which may also occur against expiration or apnoea.

Ventilation heterogeneity is multi-factorial and our observational, pragmatic study did not standardise GA at the time of study or type and degree of lung disease. It was reassuring that less mature gestational age at birth, old age and oxygen deficit were associated with greater gravity-related inhomogeneity, but not right to left inhomogeneity, even though the r^2^ values were low. Prematurity alters lung development and in early life is associated with great inhomogeneity due to surfactant deficiency.[3] In our study, infants born preterm were generally older at the time of study, and lung growth in infancy has previously been associated with increased dependent lung ventilation.[28] The finding of more ventilation inhomogeneity, but greater ventilation in the dependent lung, in those infants with greater oxygen deficit likely reflects this group of older, premature infants. These infants are likely to have more developed chest wall and diaphragmatic function than newly-born infants, as well as some degree of inflammatory parenchymal changes withouit surfactant deficiency due to evolving chronic lung injury. This highlights the utility of EIT in demonstrating biologically plausible relationships and the need for developmental and disease specific reference values for ventilation behaviour in the infant lung, an idea first proposed by Pillow, et al.[39]

## Limitations

There are a number of additional limitations to this study. There was a high imbalance between the number of synchronous (90%) and asynchronous (10%) inflations. In the only previous breath-to-breath study of synchronisation, 38% of inflations were asynchronous.[12] This likely reflects better synchronisation technology now, and suggests the ventilator used was delivering the intended therapy. The need to analyse individual inflations required a balance between feasibility and sample size. Whilst the number of infants studied was large compared to a similar study,[12] the heterogeneity was high. This is reflective of the diverse population of infants needing mechanical ventilation in the modern NICU; however, caution should be used in translating these findings into specific lung pathologies. Despite this, our results highlight the need to consider ventilation patterns based on the functional state of the lung, specifically the suitability of applied respiratory settings rather than solely the lung pathology.

There are well-described limitations of EIT, specifically the choice of image reconstruction thorax model will affect the outcome parameters.[18] As the same model was used for all analysis within this study, it does not influence comparison, but choice of model is important when referencing our findings to other EIT studies.

## Conclusions

Synchronous mechanical ventilator lung inflations generate greater gravity dependent lung ventilation and more homogenous right to left ventilation than asynchronous inflations. Even in stable infants mostly without active lung disease, regional ventilation is highly variable and creates complex lung states that may be associated with increased injury potential. Further research is warranted to determine the impact of synchronisation in mechanically ventilated infants with acute lung disease, in which ventilation heterogeneity and the risk of lung injury is greatest.

## Data Availability

Deidentified individual participant data, study protocols and statistical analysis codes are available from three months to 23 years following article publication to researchers who provide a methodologically sound proposal, with approval by an independent review committee (learned intermediary). Proposals should be directed to to gain access. Data requestors will need to sign a data access or material transfer agreement approved by MCRI.

## Abbreviations

CO_2_: Carbon Dioxide
CoV_RL_: Centre of Ventilation along the right to left lung plane
CoV_VD_: Centre of Ventilation along the ventral dorsal lung plane
EIT: Electrical Impedance Tomography
FiO_2_: Fraction of inspired oxygen
GA: Gestational Age
NICU: Neonatal Intensive Care Unit PEEP Positive end-expiratory pressure
PIP: Positive inflation pressure
ΔP: Change in pressure
ROI: Region of Interest
SpO_2_: Peripheral oxygen saturation
V_Tao_: Tidal volume at the airway opening measured by the Florian Respiratory Function Monitor
V_TEIT_: Tidal volume measured by EIT

## Acknowledgements

The authors acknowledge the infants and families involved in the study, as well as the Butterfly Ward nursing staff for their assistance in infant care during the study. The authors also acknowledge Andreas Waldmann for engineering the EIT image reconstruction infant thorax model from CT chest data provided by the Murdoch Children’s Research Institute (MCRI) Neonatal Research Group, Associate Professor Susan Donath for her advice in data analysis and Professor Inez Frerichs and Professor Colin Morley for guidance in data interpretation.

